# Experiences with the implementation of a no-kill policy for the control of feline sporotrichosis in free-roaming cats

**DOI:** 10.1101/2025.09.25.25336677

**Authors:** Raquel Geovana Nunes Alves, Blenda Araujo Martins Ferreira, Débora de Oliveira Santos, Lidiane da Silva Jesus, Bianca Moreira de Souza, Gustavo Ruas de Araújo, Graciela Kunrath Lima, Maria Isabel de Azevedo, Fernanda do Carmo Magalhães, Danielle Ferreira de Magalhães Soares, Camila Stefanie Fonseca de Oliveira

## Abstract

Sporotrichosis is an emerging zoonosis with growing impact in Brazil’s urban areas, aggravated by the absence of effective public health policies and standardized surveillance protocols, especially for free-roaming cats. This study evaluated the implementation of a surveillance and control protocol for sporotrichosis in an urban area with high densities of community cats and frequent abandonment, including animals testing positive for the disease. A retrospective, descriptive, observational epidemiological study was conducted from January 2020 to February 2025. The protocol comprised systematic monitoring, laboratory and clinical diagnostics, treatment, epidemiological surveillance, and mandatory reporting. Animals were categorized as community or abandoned. Of 24 suspected cases, 54.1% were confirmed, with 53.8% of positives originating from abandoned animals. Statistical associations were found between animal origin, reproductive status, treatment location, treatment duration, and outcomes. Abandoned cats, mainly unneutered males with multiple lesions, required approximately twice the treatment time of community cats and posed greater logistical and financial challenges. In contrast, neutered community cats with fixed feeding points and committed caregivers presented shorter treatment durations and feasible in situ therapy. As a complementary strategy, Trap-Neuter-Return (TNR) after clinical recovery was essential to reduce intra-colony transmission. The findings highlight that effective control of feline sporotrichosis depends on continuous and integrated actions aligned with the One Health approach. Individualized treatment, combined with population management and community involvement, offers an ethical and sustainable alternative to euthanasia for free-roaming cats.

## Introduction

Brazil has a population of 121.3 million dogs and cats, of which approximately 35% live on the streets [1]. These animals, in addition to the suffering they face on the streets, generate socioeconomic and environmental costs and represent a challenge for public health, as they are involved in scratching or biting accidents, predation on wildlife, and transmission of zoonotic diseases, such as sporotrichosis [2, 3].

Zoonotic sporotrichosis, transmitted mainly by cats, is caused mainly by *Sporothrix brasiliensis* in Brazil, a species recognized for its increased virulence and widespread geographic distribution [4–7]. It is a neglected subcutaneous mycosis that is widely dispersed throughout the country, with a high number of cases in the Southeast and South regions [8]. In Belo Horizonte, the capital of the state of Minas Gerais, in the southeast region, the first cases of sporotrichosis were recorded in 2015. Since then, there has been a significant increase in the incidence of the disease, which is widespread in all administrative regions of the city, as well as in neighboring municipalities [9].

Sporotrichosis is a public health problem, especially in urban regions with high population density. For its control and prevention, it is necessary to implement multidisciplinary strategies, with approaches that include actions in human, animal and environmental health [5, 10, 11]. However, despite the growing impact of the disease in Brazil, the country still lacks an effective Surveillance and Control Program for the disease, there are no comprehensive public policies or standardized protocols to monitor and contain its spread, especially in free-roaming cats populations, which live in public areas, poor communities or are abandoned in institutional spaces, including university campuses. The public service faces serious difficulties in handling feral cat colonies, there is a lack of trained people to capture and neuter them, and for these animals without guardians, the recommendation of the Municipal Programs to Combat Sporotrichosis in most of Brazil is still euthanasia.

On university campuses, the availability of resources and the presence of people willing to care for these animals make them recurrent points of abandonment, which generates conflicts within the academic community [12]. As a measure for the management of free-living animals and especially the control of zoonoses, it is necessary to institute ethical and humane population management programs that have One Health as their premise [13, 14].

The No-Kill movement, which advocates an end to the euthanasia of animals considered healthy and adoptable, began informally in the United States of America (USA) around the 1930s. However, it was only in the 1990s that this initiative gained greater visibility and strength, progressively expanding to other countries, states and municipalities on the American continent. In Brazil, the movement had its initial milestone with the enactment of Law n°4.808/2006 in the state of Rio de Janeiro, which prohibits the elimination of animals as a method of population control [14]. More recently, Federal Law n° 14.228/2021, which came into force in 2022, prohibits the elimination of dogs and cats in public establishments, except in cases of serious and incurable infectious diseases or those that pose a risk to other animals and public health [15].

Trap-Neuter-Return (TNR) is an ethical population management method widely used to control colonies of free-roaming and feral cats, and is widespread both in Brazil and in other countries [16–20], and is also applied on university campuses [12, 21]. It is a non-lethal technique, which consists of collecting, surgically sterilizing, identifying, registering, and returning the animal to the place of capture [14, 22], and may include anti-rabies vaccination and complementary tests [23, 24].

Considering the growing territorial expansion of sporotrichosis in Brazil, the increase in feral colony populations, the lack of protocols for controlling the disease and the management of these populations, this study aimed to describe and analyze the data from a program of humane ethical management of dogs and cats that proposes an alternative to euthanasia for the control of sporotrichosis in free-roaming cats.

## Materials & methods

### Type of study

A retrospective descriptive observational epidemiological study was carried out using data provided by the Permanent Animal Policy Commission of the Federal University of Minas Gerais (CPPA-UFMG) from January 2020 to February 2025. The variables analyzed included the number of positive animals, their origin (new abandonment or community, unit of belonging or sighting, species, sex, reproductive status, diagnosis, treatment time, outcome and destination.

### Study area

The study was carried out on the Pampulha campus of the Federal University of Minas Gerais (UFMG), in Belo Horizonte. The campus was founded in 1962 and has an area of 3.34 km², 50% of which is permanent preservation. It houses 22 academic units and 21 administrative units, as well as community spaces [25]. Its entrances remain accessible during opening hours, allowing animals to circulate. The extensive green area, which interfaces with surrounding roads, also makes perimeter surveillance difficult [12].

In 2018, the UFMG Rectorate set up the UFMG Permanent Animal Policy Commission (CPPA-UFMG) to promote ethical management of these populations and wildlife surveillance. The aim was to reduce birth rates, mortality, morbidity, and abandonment, as well as promote the aging of community animals, the prevention of diseases, and the control of zoonoses. To this end, the Commission implemented the TRN method, which, in addition to surgical sterilization, included testing cats for feline immunodeficiency virus (FIV) and feline leukemia (FeLV), testing dogs for visceral leishmaniasis, and anti-rabies and polyvalent vaccination.

### Management of domestic animals on the Pampulha campus - UFMG

On the Pampulha campus, CPPA-UFMG has structured a flow of surveillance and management of dogs and cats. Through face-to-face meetings, 54 people with an affinity for the animal cause were identified in 37 academic and administrative units and appointed by their directorates to be responsible for monitoring and feeding the animals in each area. Every year, a campus-wide census is carried out, with in situ observation by the appointees. Animals with ties to the academic community and frequently sighted were considered community animals and included in the TRN method, guaranteeing 100% neutering and closing the colony by 2023. All the animals were recorded in a detailed database, which includes clinical and reproductive history, date of sterilization, microchip registration, vaccinations, complementary exams and fate (return to campus, adoption, temporary home, euthanasia or natural death).

Notification of new sightings of abandoned animals was made by appointed representatives, doormen and security guards (trained to recognize resident animals) via messaging app, including a photographic record and information about the unit where the new animal was identified, gender, behavior and physical condition, for later recording in the database. The CPPA-UFMG assessed each case, checking for the presence of a microchip and whether the animal met the criteria for immediate selective collection (elderly, sick, pregnant or in heat females, infant or wild puppies), whereupon they were sent for clinical assessment, taken to the UFMG Transitional Reception and Adoption Center (CATA) and the outcome analyzed individually. Animals that didn’t meet these criteria and weren’t neutered were monitored for seven days and, if they remained on campus, they went through the TRN and were returned to their unit of origin within 15 days. Those that remained on campus were considered community animals.

### Sporotrichosis surveillance and control of esporotricose at UFMG’s Pampulha campus

Surveillance of sporotrichosis at UFMG’s Pampulha Campus is carried out according to the flow described above. The animals are monitored in situ by appointed and volunteer caretakers, as well as doorman and security guards who have received prior training. These were responsible for observing and caring for the animals on campus, reporting any new individuals or suspected lesions suggestive of sporotrichosis to the CPPA.

In suspected cases, the animals were captured/collected for confirmatory examination (mycological culture [26]). In animals that were approachable and had a closer bond with their caretakers, the collection was carried out in situ, with friendly restraint and the aid of personal protective equipment (PPE) including gloves, an apron and a mask. More skittish animals or those with larger lesions were captured using automatic traps, drop traps or forceps, always wearing PPE, and then sent to the UFMG Veterinary Hospital for sample collection with sedation when necessary. In the case of animals with a feral profile, where capture was unfeasible, the diagnosis was based on clinical and epidemiological criteria, taking into account the clinical signs observed and the environmental context. Preventive treatment was instituted and, whenever possible, the animals were kept at CATA until the results were obtained.

Samples were collected using sterile swabs, which were carefully rubbed over the selected area and then stored in tubes containing Stuart transport medium for microbiological analysis. At the same time, the swabs were imprinted onto glass slides with a frosted edge, pressing them three times at different points on the ulcerated lesion for cytological analysis. The swabs collected were processed and analyzed at LABIOMIC (Laboratory of Molecular Biology and Mycology) in the Department of Preventive Veterinary Medicine (DMVP) at the UFMG Veterinary School. The cytology slides were processed and interpreted at the Clinical Pathology Laboratory, also at the UFMG Veterinary School. All suspected or confirmed cases were documented and compulsorily notified to the Belo Horizonte Municipal Health Department, even before the obligation was established in 2024 by DAPS Joint Technical Note N° 011/2024.

Animals that tested positive for fungal culture or cytology were referred for treatment. In all cases, itraconazole was administered at a dose of 100 mg/day, or in combination with potassium iodide at a dose of 2.5 to 5 mg/kg, according to each clinical case. The medication was given once a day, in the first meal of the day, along with a small amount of moist food, such as sachets, to optimize absorption and ensure complete ingestion of the medication(s). The rest of the food was then given to each animal. The treatment was maintained until the lesions had completely healed, the hair had grown back and the respiratory signs had ceased, characterizing a clinical cure. After this phase, therapy was extended for a further 30 days to prevent recurrence. In animals with respiratory symptoms and increased nasal volume, treatment was extended for a further 60 days after the clinical cure.

The choice of treatment site took into account different factors, such as the animal’s origin, the presence of committed and available carers, the existence of a feeding point, the daily frequency of food provision (Monday to Monday, without an automatic dispenser at the weekend) and the location in areas of risk to the academic community (for example near children’s schools, restaurants or other places with a greater flow of people) (Figure 1). The treatment was therefore divided into two groups:

1. Animals suspected of having sporotrichosis that were abandoned or in the community where it was difficult to standardize feeding or had comorbidities were sent to temporary homes with an isolation area and caregivers trained to administer medication, reducing risks. Preferably, these animals were treated in partner veterinary clinics that had adequate facilities, which also applied to community animals without regular daily sightings or a caregiver responsible for feeding.
2. On the other hand, community animals with established feeding points, with caretakers who carried out daily management and a guaranteed sighting frequency were treated in situ. In these cases, there was daily notification to CPPA-UFMG with photos of the animal to monitor clinical progression and empty feeders after administration of the medication.

**Figure 1.**
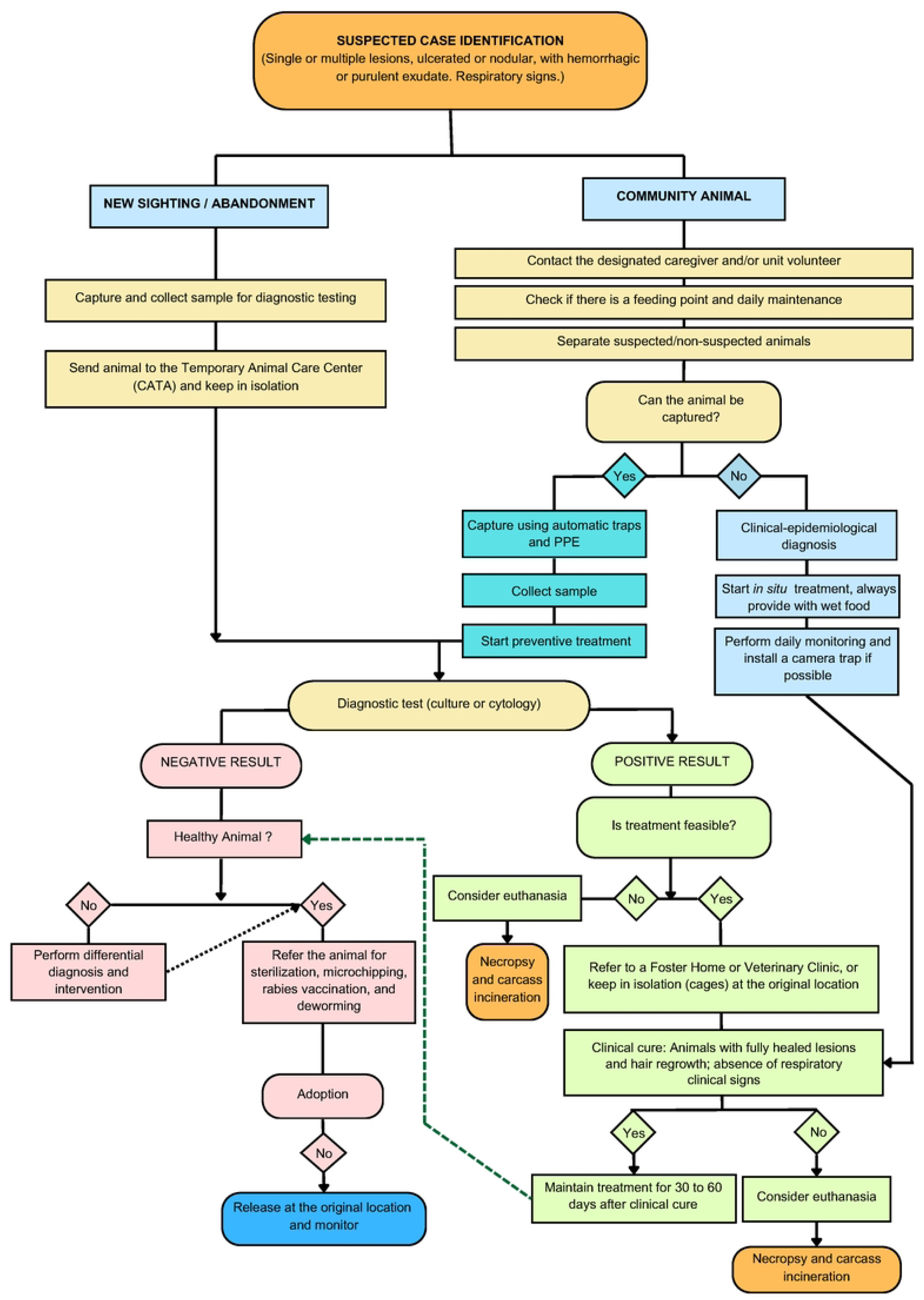
Flowchart of treatment of animals positive for sporotrichosis at the Pampulha Campus by CPPA-UFMG, 2022 to 2025.

The outcomes of the cases followed two main scenarios. In the first, animals that showed remission of wounds and clinical healing and were not neutered were sent to the TRN. Animals whose origin was abandonment were preferably put up for adoption, while those not adopted or those already living on campus (community animals) were returned to their unit of origin. In the second scenario, those who showed resistance to treatment or concomitant health conditions that hindered their recovery were euthanized. The carcasses were sent for necropsy and then incinerated by a third-party company from the UFMG Veterinary Hospital. All the people involved in caring for suspected animals, as well as security guards and doormen, received guidance on zoonotic sporotrichosis and the preventative measures that should be taken to avoid infection.

### Statistical analysis

The animals were allocated into two groups according to their origin: abandoned (new animal) or community (resident), for comparison purposes. The data was tabulated and subjected to frequency distribution analysis. Fisher’s exact test or Pearson’s chi-squared test was performed to verify associations between categorical variables, adopting a significance level of 5% (p-value significant when ≤ 0.05), followed by analysis of Pearson’s standardized residuals, considering an association significant when the residual value was greater than 1.96. All the analyses were carried out using Stata/MP software version 16.0. The analysis of the spatial distribution of cases was carried out using QGIS software, version 3.42.1 and the cartographic bases used were downloaded from the pages of the Brazilian Institute of Geography and Statistics (IBGE) where they are freely available.

### Ethical statement

The study protocol was approved by the Ethics Committee on the Use of Animals of the Federal University of Minas Gerais (CEUA/UFMG) under number 60/2022, approved on June 6, 2022 (valid until June 5, 2027).

## Results

### Origin and spatial distribution of suspected and confirmed sporotrichosis animals

Between January 2020 and January 2025, 24 animals with suspected sporotrichosis lesions were reported in the study area, 12 (50.00%) of which were new (abandoned) animals and 12 (50.00%) community (resident) animals in 14 (32.6%) of the 43 units (Figure 2).

**Figure 2.**
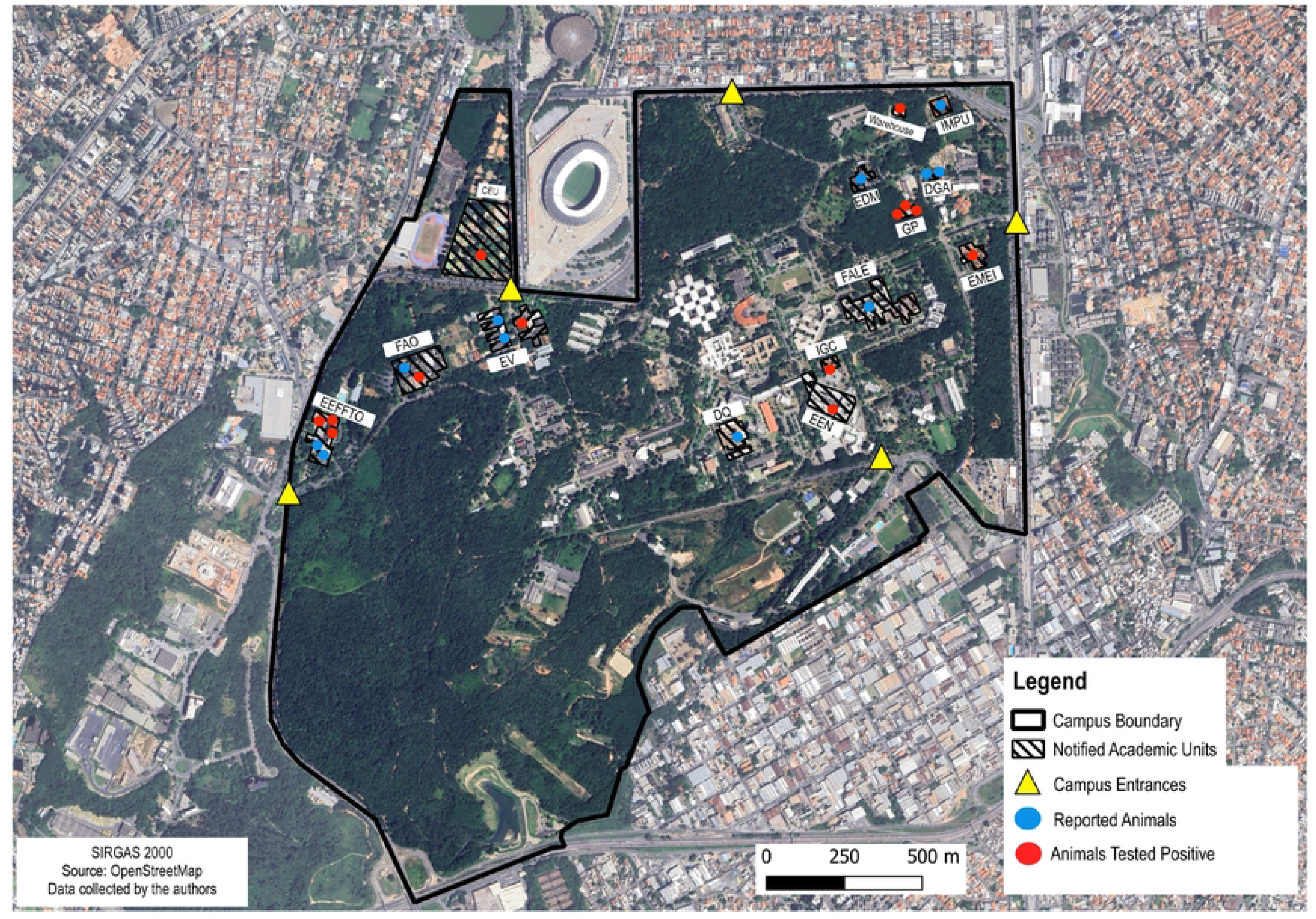
Distribution of Notified and Positive Cases on the Pampulha Campus - UFMG (2020-2025).

There was an association between academic units and the occurrence of abandoned animals with suspected sporotrichosis (p=0.014, 95% CI). The unit called the School of Physical Education, Physiotherapy and Occupational Therapy (EEFFTO) was the only unit in which there was an association with abandoned animals. Table 1 shows the absolute and relative frequency of notifications, as well as the association analyses by academic unit.

**Table 1.** Distribution and association between Academic Units and abandoned and community animals with suspected ichosis on the Pampulha Campus - UFMG (2020-2025)

| Academic Unit | Abandoned (OF) | Abandoned (RF) | Abandoned (AR) | Community (OF) | Community (RF) | Community (AR) |
| --- | --- | --- | --- | --- | --- | --- |
| Almoxarifado | 0 | 0.00% | -1.022 | 1 | 4.17% | 1.022 |
| CEU | 0 | 0.00% | -1.022 | 1 | 4.17% | 1.022 |
| DGA | 1 | 4.17% | 0.000 | 1 | 4.17% | 0.000 |
| DQ | 1 | 4.17% | 1.022 | 0 | 0.00% | -1.022 |
| EDM | 1 | 4.17% | 1.022 | 0 | 0.00% | -1.022 |
| EEFFTO | 5 | 20.83% | 2.513 | 0 | 0.00% | -2.513 |
| EEN | 1 | 4.17% | 1.022 | 0 | 0.00% | -1.022 |
| EMEI | 1 | 4.17% | 1.022 | 0 | 0.00% | -1.022 |
| EV | 0 | 0.00% | -1.852 | 3 | 12.50% | 1.852 |
| FAFICH | 0 | 0.00% | -1.022 | 1 | 4.17% | 1.022 |
| FAO | 0 | 0.00% | -1.477 | 2 | 8.33% | 1.477 |
| GP | 1 | 4.17% | -0.617 | 2 | 8.33% | 0.617 |
| IGC | 1 | 4.17% | 1.022 | 0 | 0.00% | -1.022 |
| IMPU | 0 | 0.00% | -1.022 | 1 | 4.17% | 1.022 |
| TOTAL | 12 | 50.00% | - | 12 | 50.00% | - |

*Note:* OF = Observed frequency (actual number of cases); RF = Relative frequency (number of cases in relation to the total notifications); AR = Adjusted residual (indicates the deviation from expected frequency, considered significant when above $\pm 1.96$ ).
CEU (University Sports Center), DGA (Environmental Management Department), DQ (Department of Chemistry), EDM (School of Music), EEFETO (School of Physical Education, Physiotherapy and Occupational Therapy), EEN (School of Engineering), EMEI (Alaíde Lisboa Municipal Early Childhood Education School), EV (School of Veterinary Medicine), FAFICH (Faculty of Philosophy and Human Sciences), FAO (School of Dentistry), GP (Forte Warehouse), IGC (Institute of Geosciences), and IMPU (University Press).

A comparison of the frequencies of the clinical, treatment and resolvability variables according to group is shown in Table 2.

**Table 2.** Distribution of animals suspected of sporotrichosis according to clinical variables, treatment and resolvability at the Pampulha Campus - UFMG (2020-2025)

| Variable | Abandoned |  | Community |  | Total |  | *P-value* |
| --- | --- | --- | --- | --- | --- | --- | --- |
|  | N | % | N | % | N | % |  |
| Species |  |  |  |  |  |  |  |
| Feline | 11 | 45.83% | 12 | 50.00% | 23 | 95.83% | 1.000 |
| Canine | 1 | 4.17% | 0 | 0.00% | 1 | 4.17% | 1.000 |
| Sex |  |  |  |  |  |  |  |
| Female | 5 | 20.83% | 5 | 20.83% | 10 | 41.67% | 1.000 |
| Male | 7 | 29.17% | 7 | 29.17% | 14 | 58.33% | 1.000 |
| Reproductive Status * |  |  |  |  |  |  |  |
| Not neutered | 10 | 41.67% | 0 | 0.00% | 12 | 50.00% | 0.000 |
| Neutered | 2 | 8.33% | 12 | 50.00% | 12 | 50.00% | 0.000 |
| Lesion Location |  |  |  |  |  |  |  |
| Ear | 2 | 8.33% | 2 | 8.33% | 4 | 16.67% | 0.329 |
| Face | 1 | 4.17% | 4 | 16.67% | 5 | 20.83% | 0.329 |
| Snout | 3 | 12.50% | 1 | 4.17% | 4 | 16.67% | 0.329 |
| Limbs | 1 | 4.17% | 4 | 16.67% | 5 | 20.83% | 0.329 |
| Not reported | 4 | 16.67% | 2 | 8.33% | 6 | 25.00% | 0.329 |
| Capture Status |  |  |  |  |  |  |  |
| Captured | 10 | 45.45% | 11 | 50.00% | 21 | 87.50% | 1.000 |
| Not captured | 0 | 0.00% | 1 | 4.55% | 1 | 4.17% | 1.000 |
| Natural death | 1 | 4.55% | 1 | 4.55% | 2 | 8.33% | 1.000 |
| Initial Prognosis * |  |  |  |  |  |  |  |
| Favorable | 2 | 8.33% | 9 | 37.50% | 11 | 45.83% | 0.017 |
| Guarded | 2 | 8.33% | 0 | 0.00% | 2 | 8.33% | 0.017 |
| Unfavorable | 4 | 16.67% | 1 | 4.17% | 5 | 20.83% | 0.017 |
| Not reported | 4 | 16.67% | 2 | 8.33% | 6 | 25.00% | 0.017 |
| Cytology Result |  |  |  |  |  |  |  |
| Positive | 2 | 8.33% | 0 | 0.00% | 2 | 8.33% | 0.549 |
| Negative | 2 | 8.33% | 1 | 4.17% | 3 | 12.50% | 0.549 |
| Inconclusive | 1 | 4.17% | 0 | 0.00% | 1 | 4.17% | 0.549 |
| Not performed | 7 | 29.17% | 11 | 45.83% | 18 | 75.00% | 0.549 |
| Culture Result |  |  |  |  |  |  |  |
| Positive | 6 | 25.00% | 3 | 12.50% | 9 | 37.50% | 0.370 |
| Negative | 5 | 20.83% | 6 | 25.00% | 11 | 45.83% | 0.370 |
| Not performed | 1 | 4.17% | 3 | 12.50% | 4 | 16.67% | 0.370 |
| Treatment Status<sup>a</sup> |  |  |  |  |  |  |  |
| Yes | 6 | 25.00% | 5 | 20.83% | 11 | 45.83% | 1.000 |
| No | 2 | 8.33% | 1 | 4.17% | 3 | 12.50% | 1.000 |
| Negative | 5 | 20.83% | 5 | 20.83% | 10 | 41.67% | 1.000 |

| Treatment Location* |  |  |  |  |  |  |  |
| --- | --- | --- | --- | --- | --- | --- | --- |
| Partner clinic | 3 | 12.50% | 0 | 0.00% | 3 | 12.50% | 0.004 |
| Temporary home | 3 | 12.50% | 0 | 0.00% | 3 | 12.50% | 0.004 |
| In situ | 0 | 0.00% | 5 | 20.83% | 5 | 20.83% | 0.004 |
| Not performed | 6 | 25.00% | 7 | 29.17% | 13 | 54.17% | 0.004 |
| Outcome |  |  |  |  |  |  |  |
| Clinical cure | 4 | 16.67% | 2 | 8.33% | 6 | 25.00% | 0.307 |
| Relapse | 0 | 0.00% | 1 | 4.17% | 1 | 4.17% | 0.307 |
| Treatment suspension | 0 | 0.00% | 2 | 8.33% | 2 | 8.33% | 0.307 |
| Euthanasia | 1 | 4.17% | 0 | 0.00% | 1 | 4.17% | 0.307 |
| Natural death | 1 | 4.17% | 0 | 0.00% | 1 | 4.17% | 0.307 |
| Negative | 6 | 25.00% | 7 | 29.17% | 13 | 54.17% | 0.307 |
| Destination |  |  |  |  |  |  |  |
| Adoption | 8 | 33.33% | 2 | 8.33% | 10 | 41.67% | 0.003 |
| Return to the origin | 0 | 0.00% | 7 | 29.17% | 7 | 29.17% | 0.003 |
| Natural death | 3 | 12.50% | 1 | 4.17% | 4 | 16.67% | 0.003 |
| Not reported | 2 | 8.33% | 1 | 4.17% | 3 | 12.50% | 0.003 |

\* Statistical significance was determined using Fisher's exact test.
<sup>a</sup>Animals under preventive treatment and positive based on laboratory and clinical-epidemiological criteria.

### Criteria for confirming the disease

Of the 24 suspected animals, confirmatory tests (culture and/or cytology) were carried out on 83.33% (20/24) of the individuals. Preventive treatment was started in 45.83% (11/24) and suspended after a negative confirmatory test result. In total, 54.16% (13/24) of the animals were diagnosed as positive, of which 11 were based on laboratory criteria (84.62%) and only 15.38% (2/13) were confirmed based on clinical-epidemiological criteria.

### Species, sex, reproductive status and origin of the positive animals

Most of the positive cases were cats (92.30%, 12/13), while only one (7.69%, 1/13) was a dog. In terms of sex, 81.53% (8/13) were male and 38.46% (5/13) female and 81.53% (8/13) were neutered. Among the positive animals, 53.84% (7/13) were abandoned (animals new to the campus) and 46.15% (6/13) were community animals (residents).

### Treatment for sporotrichosis and the outcome of cases

Among the positive animals, 76.92% (10/13) were treated for sporotrichosis and 23.07% (3/13) were not treated due to the presence of comorbidities and an unfavorable prognosis. As for the fate of the positive animals, 38.46% (5/13) were returned to their place of origin on campus and 30.76% (4/13) were donated, 15.38% (2/13) were euthanized and 15.38% (2/13) died before treatment was instituted, as they were abandoned on campus in an advanced stage of the disease. The animals that died were sent for necropsy, followed by incineration of the carcasses.

Some variables showed a significant association. All the community animals (residents) were neutered (p-value = 0.000). The prognosis of community animals was more favorable than that of abandoned animals (p-value=0.012). With regard to the treatment setting, community animals were predominantly treated *in situ* (p-value=0.012). In addition, the average treatment time was significantly longer for abandoned animals (p-value=0.0357), being approximately twice as long as for community animals.

As shown in Figure 3, the community animals were treated for an average of 2.2 months, while the abandoned animals had an average treatment time of 4.8 months.

**Figure 3.**
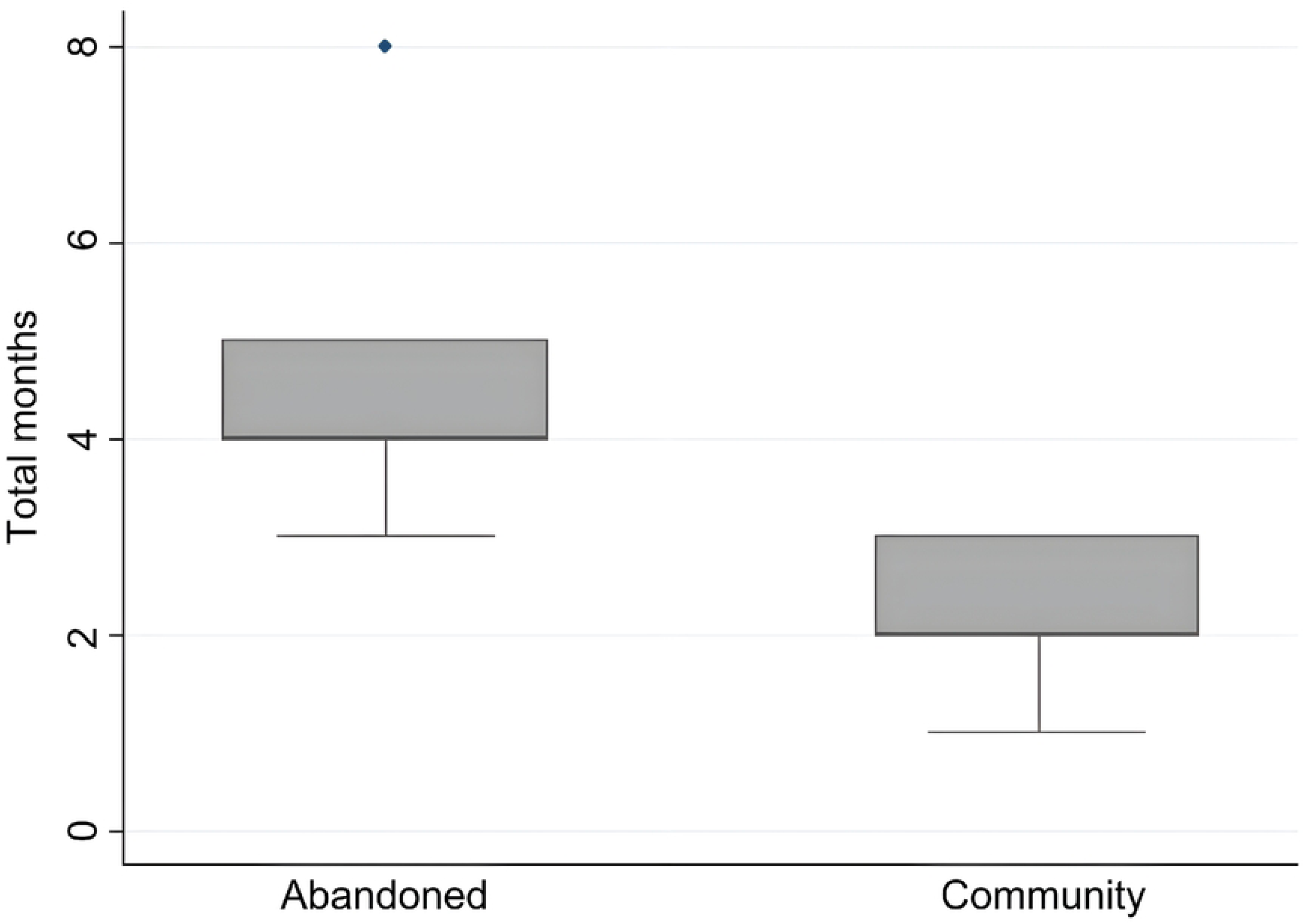
Treatment time in months of abandoned and community animals positive for sporotrichosis on the Pampulha Campus - UFMG (2020-2025)

The final destination of the animals also differed between the groups (p-value = 0.013): while the main destination for abandoned animals was adoption (residual value = 2.083), community animals were mostly returned to their place of origin (residual value = 3.121), as shown in Figure 4.

**Figure 4.**
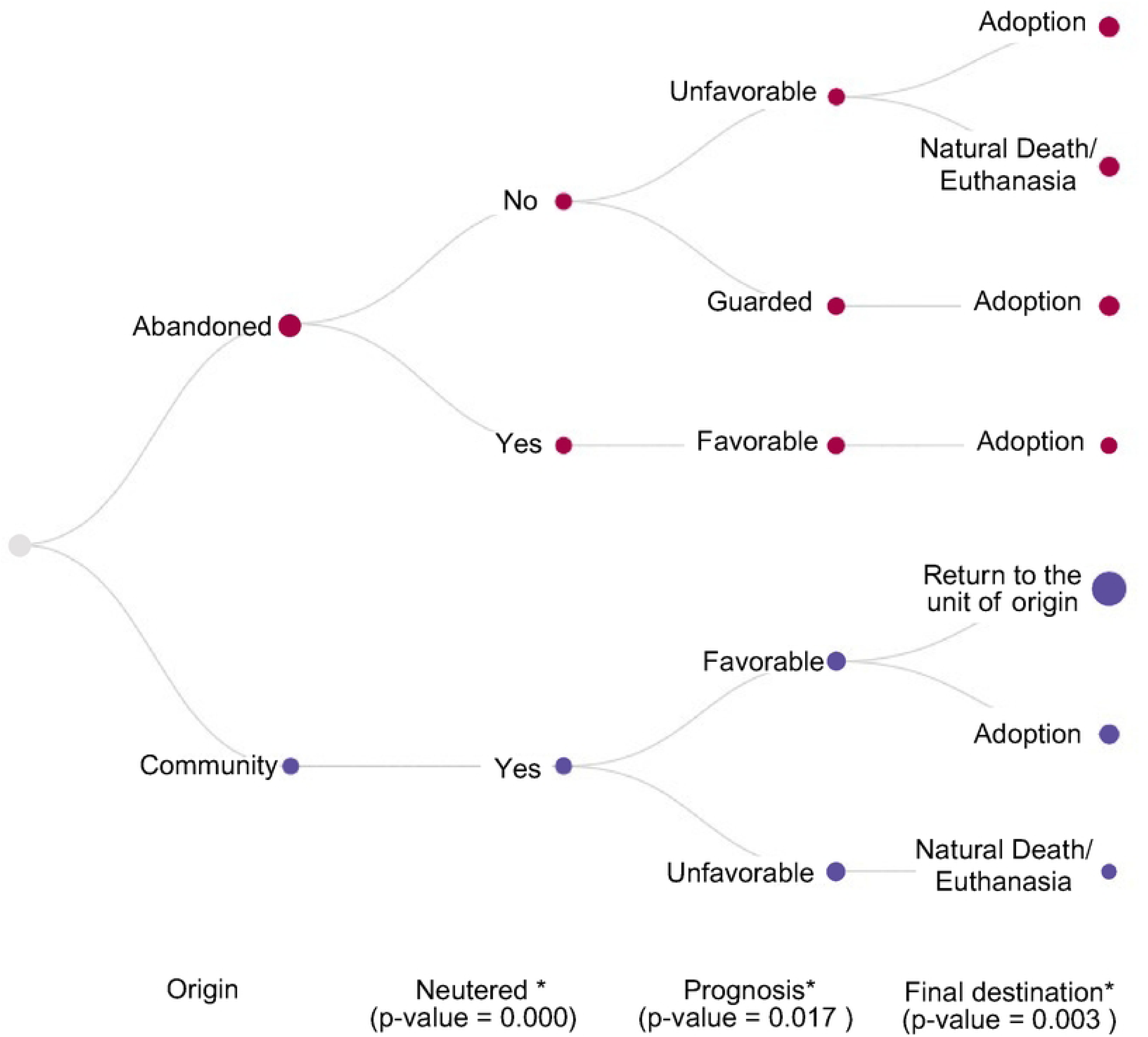
Hierarchy map of comparison between groups of the variables reproductive status, prognosis and destination of animals suspected of sporotrichosis on the Pampulha Campus - UFMG (2020-2025). *Statistical significance was determined using Fisher’s exact test (p-value ≤ 0.05)

There was no significant association between the variables species, sex, location of the lesion, capture, positivity, criteria for confirming the diagnosis (cytology, culture and clinical-epidemiological), institution of treatment, therapeutic protocol (itraconazole in monotherapy or in combination) and outcome (clinical cure, suspension of treatment, death, euthanasia and relapse), as verified by Fisher’s Exact Test.

## Discussion

CPPA-UFMG has set up a monitoring and surveillance network, as well as a solid population management program over the last five years, with stabilization of the resident (community) animal population. Despite this, UFMG still has high abandonment rates [12]. The gates have no entry control during university hours, which allows free movement on campus and corroborates the migration and abandonment of animals.

This scenario may explain why the unit with the highest number of animals suspected of sporotrichosis was the EEFFTO, as it is located at one of the university’s main entrances and lacks constant surveillance, making it easier for animals to be abandoned.

Most of the positive animals were cats and males. These findings are consistent with the cases described in the literature, whose main risk factors for zoonotic sporotrichosis include felines with access to the street, males, not neutered and involved in disputes over territory and females, increasing the likelihood of infection [26–29]. In the present study, the castration variable differed from the literature because most of the animals had already undergone CED and were fixed. However, the positivity can still be explained by the fact that these animals, in the case of community animals, lead a free life, roaming in open areas, scratching tree trunks [5] and other behavioral factors, such as disputes over food.

For diagnosis, most cases were confirmed by laboratory criteria, using cytology and fungal culture, the latter being considered the gold standard test [28, 30]. In some cases, when it has not been possible to collect material, clinical epidemiological diagnosis has been chosen, considering that it is a suspected case that has an epidemiological link with other confirmed animals and in an area with known transmission. Although not ideal, according to the Ministry of Health, it can be used in endemic areas to prevent the spread of the disease [8].

As for the outcome of the cases, when comparing the groups, although the community animals had a better initial prognosis than the abandoned animals, there was no difference in the percentage of clinical cures. Although there was no correlation in this study between the therapy instituted and the cure, studies have shown that monotherapy with Itraconazole has cure rates varying from 38.3% to 100% [31, 32] and that in association with Potassium Iodide the cure rate increases to 88% to 96.15% of cases with an average of 3.5 months of treatment [4, 29]. The average treatment time is between 4 and 9 months, and it is necessary to continue the medication after clinical cure [33]. Medication should be maintained for at least one month after remission of clinical signs, and for two months in cases where the respiratory system is involved, to reduce the risk of relapse [5]. In the cases evaluated, the treatment time for each animal varied between one and six months, reflecting the differences in responses to treatment, which are influenced by factors such as the severity of the infection and the individual response of each animal [33]. However, in the group of community animals, it was found that the cure was achieved in half the time when compared to the cure in abandoned animals.

When the fate of the animals after diagnosis was analyzed, the findings of this study showed that in situ treatment exclusively for resident (community) animals can be an alternative for public health purposes and an alternative to euthanasia for free-living animals, since the treatment time for these animals was shorter than for abandoned individuals and there were no reports of human cases or new animals in the respective animal units. It is extremely important to closely monitor each animal treated in the field, administering the drug in a small amount of food, in the first meal of the day, ensuring complete ingestion and only after giving the rest of the food. Photographing the animals every day to follow their clinical progress and monitoring the other animals in the colony with images are essential. Otherwise, difficulties in confinement can result in irregularities in the administration of medication, which can lead to a recurrence of the disease, hindering the healing process and favoring the spread of sporotrichosis [33, 34].

The absence of new feline cases in the treated units or human cases shows that the establishment of an effective and viable therapeutic protocol for cats in field conditions is essential for the control of zoonotic sporotrichosis [5]. However, for this protocol to be applied, criteria must be taken into account that depend directly on the commitment of the caregiver and the monitoring of trained professionals, avoiding therapeutic errors and recurrence of the disease [35]. The existence of a fixed feeding point and daily maintenance is fundamental to the success of the action since itraconazole and potassium iodide capsules can be opened and administered with wet food, which facilitates absorption and increases the bioavailability of the drug, as well as reducing the risk of zoonotic transmission because there is no direct contact with the positive animal [4, 5]. In places where there are no caregivers to ensure this, the animals should be collected, especially in cases of abandonment. Volunteer caregivers should receive training on the disease, understanding the risks, prevention measures and the importance of committing to treatment.

According to de Souza et al. [36], when health education actions were implemented for guardians of cats affected by sporotrichosis, treatment time was reduced from 131.2 days to 67.5 days, a reduction of 48.55%. These findings highlight the relevance of educational strategies as tools to improve therapeutic efficacy, reducing both treatment time and the risk of abandonment, as well as contributing to the control of sporotrichosis transmission in the context of single health.

As an associated control and prevention measure, subjecting free-living animals to NRT after clinical cure is essential to reduce the chances of transmission within the colony. Ideally, preventing animals from freely roaming the streets is only feasible through responsible adoption, which in Brazil is not yet a reality due to the high number of animals for few available homes, despite the significant allocation of abandoned animals for adoption observed in the study. For community animals, the return to the unit of origin, combined with continuous monitoring by the community involved, as well as the use of tools such as camera traps for surveillance, is essential to identify possible relapses.

The surveillance and control of sporotrichosis in free-living cats is a constant challenge for public health. Actions need to be continuous, integrated and guided by the One Health concept. The availability of financial, physical and human resources is a determining factor in the creation of public policies and effective guidelines for the control of zoonotic sporotrichosis. In this context, ethical population management programs are fundamental to reducing abandonment and controlling stray populations, as demonstrated by the work of CPPA-UFMG on the Pampulha campus.

The results reinforce the need for an active surveillance network to identify suspected cases early and refer them for diagnosis. In addition, there are viable alternatives to euthanizing feral cats with sporotrichosis, but their implementation depends on the structure and resources available.

## Conclusion

Capture, immediate initiation of treatment, sterilization after clinical cure and continuous monitoring are essential strategies in the management of sporotrichosis within a No-kill policy. To tackle sporotrichosis effectively, especially in free-living animals, it is not enough to treat individual cases. A collective, preventive and sustainable effort is needed, involving the entire local community, so that a single healthy and harmonious human-animal relationship can be guaranteed.

*In situ* treatment of free-roaming cats, although not the best option, maybe the only alternative when capture is unfeasible. To do this, it is essential to understand the population dynamics of the area and assess the risks posed to human and animal health by the permanence of positive animals.

The responsible caretaker must be committed to feeding and administering medication, as well as receiving information about the disease and prevention measures. Ideally, animals should be isolated to avoid environmental contamination and infection of other animals and humans. In areas with a history of sporotrichosis and positive colonies, preventive treatment even before diagnostic confirmation is essential. There is also a clear need to reinforce educational actions on responsible guardianship and combating abandonment since animals that receive proper care have a shorter treatment time and consequently a better prognosis.

The group of abandoned animals that tested positive for sporotrichosis were male cats, not neutered, with multiple lesions and a poor prognosis. This led to a treatment time twice as long, with the need to capture them and send them to clinics or temporary homes, which increased the cost. As for the group of resident animals infected with sporotrichosis, the majority were neutered animals with few lesions who were treated *in situ* with the help of local caretakers. For these animals, the cure time was halved, with a favorable prognosis, and they were returned to the campus after a complete cure.

## Acknowledgments

The authors specifically thank all the volunteers of the Dog and Cat Management Program at the Federal University of Minas Gerais (UFMG).

## Supporting information

**S1 Table.** Factors associated with the frequency of cases treated for sporotrichosis among domestic cats at UFMG from 2020 to 2025.

**S2 Table.** Number of animals attended and diagnosed for sporotrichosis on the Pampulha Campus - UFMG (2020-2025).

**S1 Fig.** Flowchart of treatment of animals positive for sporotrichosis at the Pampulha Campus by CPPA-UFMG, 2022 to 2025.

**S2 Fig.** Distribution of Notified and Positive Cases on the Pampulha Campus - UFMG (2020-2025).

**S3 Fig.** Treatment time in months of abandoned and community animals positive for sporotrichosis on the Pampulha Campus - UFMG (2020-2025).

**S4 Fig.** Hierarchy map of comparison between groups of the variables reproductive status, prognosis and destination of animals suspected of sporotrichosis on the Pampulha Campus - UFMG (2020-2025).

## Data availability statement

The original contributions presented in the study are included in the article/supplementary material, further inquiries can be directed to the corresponding author.

